## Supporting information for "Transmission dynamics and vaccination strategies for Crimean-Congo haemorrhagic fever virus in Afghanistan: a modelling study"

#### 1. Model structure

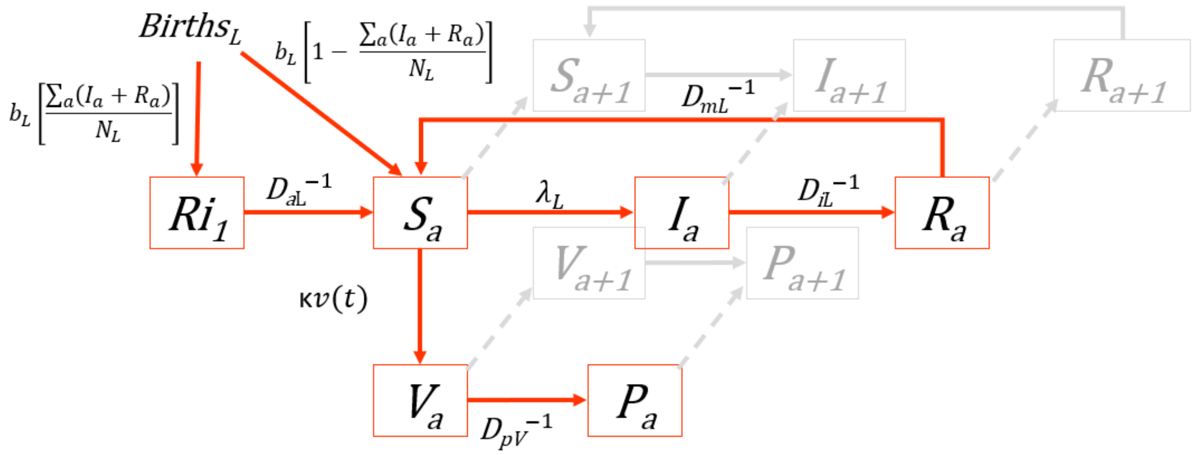

**Figure S1: Model structure for the transmission of CCHFV amongst livestock.** The structure shows the compartments and state transitions for livestock . Births occur at a rate equal to the mortality rate to maintain a population at equilibrium. Mortality rates are estimated to achieve a known livestock age-distributed population (see livestock demographic model in section 3). Offspring from prevalent CCHFV animals acquire transient immunity at birth through first colostrum ($R_i$). This immunity lasts for an assumed average period of 6 months. After this period, livestock move to the susceptible stage ($S_1$). Susceptible livestock ($S_a$) acquire CCHFV with a force of infection $\lambda_L$ that leads to an infectious period ($I_a$) with mean duration $D_{iL}$, expressed as inverse time rate $D_{iL}{}^{-1}$. We assume that livestock lose immunity at a rate $D_{mL}$.. Vaccination is implemented by recruiting susceptible animals at a rate *v(t).* The effective number of immunised livestock is also defined by the efficacy of the vaccine (κ). At first, vaccination does not confer immunity $V_a$, which is only acquired after a period of length $D_{pV}$. In this structure, subscript *a* points to the age category within the age structure, over which transitions occurs as depicted with the shaded grey structure in the background.

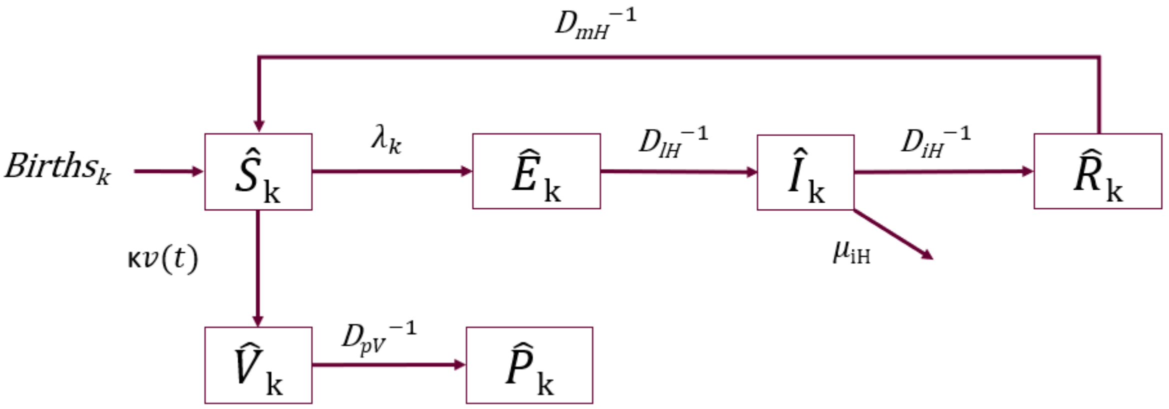

**Figure S2: Model structure for the spillover transmission of CCHFV and disease progression in humans.** Humans are born at a rate reflecting the life expectancy in Afghanistan (keeping population size constant in the absence of infections), and split into the two human categories considered in this model. Namely farmers (the high-risk group) and other occupations. This distribution is taken from previous USAID surveys in the country (see parameters table in the main text). The categorisation by occupation in the model is reflected in this structure and in the mathematical equations using subscript *k* (0= farmer; 1=others). Infection is acquired in humans with force of infection $\lambda_k$, with differential risk *k.* Infection is followed by a latent period $\hat{E}_k$ with mean duration $D_{lH}$ that leads to an infectious period $\hat{I}_k$ . This infectious period can lead to either recovery $\hat{R}_k$ or death. Death from CCHFV in humans is described in the model as the competing hazard $\mu_{iH}$ that summarise the case fatality ratio for CCHFV. We assume waning immunity in humans that leads back to susceptible stage at a rate $D_{mH}{}^{-1}$. Vaccination occurs at rate ***v(t)***, differential by occupation. Effective number of immunised people is finally defined by the efficacy of the vaccine (κ). Vaccine protection occurs after vaccination after a mean period $D_{pV}$.

### 2. Model equations

#### 2.1. Livestock model

Transmission of CCHFV among livestock is described with a compartmental deterministic model with mathematical expressions written below (equations 1 to 6):

<u>Susceptible</u>

$$\frac{dS_a(t)}{dt} = \left\{ b_L \left( 1 - \frac{\sum_a \left( I_a(t) + R_a(t) \right)}{N_L(t)} \right) + Ri(t) D_{aL}^{-1} + R_a(t) D_{mL}^{-1} - S_a(t)\left(\lambda_L(t) + \kappa v(t) + \mu_a\right) + \sum_{i=1}^{5} Z_{i,a} S_i \, \delta for a = 1 R_a(t)(D_{mL}|{-1}) - S_a(t)(\lambda L(t) + (\kappa) v(t) + \mu_a) + \sum_{i=1}^{5} Z_{i,a} S_i \, \delta for a > 1 \right.$$

*(Eq.1)*

<u>Transient colostrum immunity</u>

$$\frac{dRi(t)}{dt} = b_L \left( \frac{\sum_a I_a(t)}{N_L(t)} \right) - Ri(t) D_{aL}^{-1} - Ri(t)\mu_a$$

*(Eq.2)*

Infectious livestock

$$\frac{dI_a(t)}{dt} = S_a(t)\lambda L(t) - I_a(t)\left(D_{iL}{}^{-1} + \mu_a\right) + \sum_{i=1}^{5} Z_{i,a} I_i\,\delta$$

*(Eq.3)*

Recovered

$$\frac{dR_a(t)}{dt} = I_a(t)\left(D_{iL}{}^{-1}\right) - R_a(t)\left(D_{mL}{}^{-1} + \mu_a\right) + \sum_{i=1}^{5} Z_{i,a} R_i\,\delta$$

*(Eq.4)*

Newly vaccinated

$$\frac{dV_a(t)}{dt} = S_a(t)\kappa v(t) - V_a(t)\left(D_{pV}{}^{-1} + \mu_a\right) + \sum_{i=1}^{5} Z_{i,a} V_i\,\delta$$

*(Eq.5)*

Vaccine immunity

$$\frac{dP_a(t)}{dt} = V_a(t) D_{pV}{}^{-1} - P_a(t)(\mu_a) + \sum_{i=1}^{5} Z_{i,a} P_i\,\delta$$

*(Eq.6)*

Age transition identity matrix

$$Z_{i,j} = \begin{bmatrix} -1 & 0 & 0 & 0 & 0 \\ 1 & -1 & 0 & 0 & 0 \\ 0 & 1 & -1 & 0 & 0 \\ 0 & 0 & 1 & -1 & 0 \\ 0 & 0 & 0 & 1 & -1 \end{bmatrix} ,\textit{for } j = 1,\ldots,5, \textit{ and } i = 1\ldots5$$

*(Eq.7)*

Force of infection in Livestock

$$\lambda_L = \beta_L \frac{\sum_a I_a}{N_L}$$

*(Eq.8)*

With environmental-driver-dependent transmission probability per-capita

$$\beta_L = \frac{R_L(t)}{D_{iL}}$$

*(Eq.9)*

Where $R_L(t)$ is the reproduction number at each point in time *t.*

$R_L(t)$ is defined according to the environmental driver used in the model. In section 2.3 of this document we describe the conditions set for each driver. As mentioned in the main text, we use environmental drivers as a proxy for tick-activity. Hence, we incorporate to our best knowledge, how this drivers might affect such activity.

#### 2.2. Human spillover model

CCHFV transmission into humans in the model occurs as a function of the prevalence of CCHFV in livestock and a calibrated risk factor for farmers and non-farmers which implies the intensity of contact with animals and also the differential excess risk in farmers relative to other occupations. Here we assume that transmission into humans occurs only as a result of contact with animals and not with infectious humans. Contact with animals cover at least two potential routes: contact with animal fluids, and tick bites from ticks feeding in infectious livestock.

This spillover event is likely to be subject to stochastic variations, therefore we write a spillover model for humans as a discrete compartmental stochastic model, that follows a SEIRS structure (see Fig 1 in main text and Fig S2). Model states and transitions described below.

<u>Susceptible humans</u>

$$\begin{aligned}\hat{S}_k(t+1) = \hat{S}_k(t) + Binomial\left[b_H, \hat{N}_k(t)\right] + Binomial\left[\left({D_{mH}}^{-1}\right), \hat{R}_k(t)\right] \\ - Binomial\left[\lambda_k(t), \hat{S}_k(t)\right] - Binomial\left[\kappa v(t), \hat{S}_k(t)\right] - Binomial\left[\mu_H, \hat{S}_k(t)\right]\end{aligned}$$

*(Eq.10)*

<u>Exposed humans</u>

$$\begin{aligned}\hat{E}_k(t+1) = \hat{E}_k(t) + Binomial\left[\lambda_k(t), \hat{S}_k(t)\right] - Binomial\left[{{D_{lH}}^{-1}}^{-1}, \hat{E}_k(t)\right] \\ - Binomial\left[\mu_H, \hat{E}_k(t)\right]\end{aligned}$$

*(Eq.11)*

<u>Infectious humans</u>

$$\begin{aligned}\hat{I}_k(t+1) = \hat{I}_k(t) + Binomial\left[{{D_{lH}}^{-1}}^{-1}, \hat{E}_k(t)\right] \\ - Binomial\left[\frac{{D_{iH}}^{-1}}{\left(\mu_{iH} + {D_{iH}}^{-1}\right)}, totalEvents_k(t)\right] \\ - Binomial\left[\frac{\mu_{iH}}{\left(\mu_{iH} + {D_{iH}}^{-1}\right)}, totalEvents_k(t)\right] - Binomial\left[\mu_H, \hat{I}_k(t)\right]\end{aligned}$$

*(Eq.12)*

Recovered humans

$$\hat{R}_k(t+1) = \hat{R}_k(t) + Binomial\left[\frac{D_{iH}{}^{-1}}{\left(\mu_{iH} + D_{iH}{}^{-1}\right)}, totalEvents_k(t)\right] - Binomial\left[\left(D_{mH}{}^{-1}\right), \hat{R}_k(t)\right] - Binomial\left[\mu_H, \hat{R}_k(t)\right]$$

*(Eq.13)*

Newly vaccinated humans

$$\hat{V}_k(t+1) = \hat{V}_k(t) + Binomial\left[\kappa v(t), \hat{S}_k(t)\right] - Binomial\left[D_{pV}{}^{-1}, \hat{V}_k(t)\right] - Binomial\left[\mu_H, \hat{V}_k(t)\right]$$

*(Eq.14)*

Newly vaccine protected humans

$$\hat{P}_k(t+1) = \hat{P}_k(t) + Binomial\left[D_{pV}{}^{-1}, \hat{V}_k(t)\right] - Binomial\left[\mu_H, \hat{P}_k(t)\right]$$

*(Eq.15)*

Force of infection in humans

Force of infections in human is here a function of the prevalence of infectious livestock at time *t* and a transmission probability $\beta_F$. For other occupations we include a factor O that is estimated to reflect the risk ration between farmers and other human groups.

$$\lambda_k(t) = \begin{cases} \beta_F \dfrac{\sum_a I_a(t)}{N_L(t)} \quad , for\ farmers(k=1) \\ O\beta_F \dfrac{\sum_a I_a(t)}{N_L(t)}, for\ others(k=2) \end{cases}$$

*(Eq.16)*

To estimate new fatalities and recoveries we use the competing hazard formula, relying on the known CFR for CCHFV(see table 1 in main text). We write the corresponding competing mortality hazard as follows:

$$\mu_{iH} = \frac{CFR_{cchfv}\left(D_{iH}{}^{-1}\right)}{1 - CFR_{cchfv}}$$

*(Eq.17)*

Then we calculate the total number of events (new fatalities and recoveries) occurring at time *t* in the infectious compartment:

$$totalEvents_k(t) = Binomial\left[\left(D_{iH}{}^{-1} + \mu_{iH}\right), \hat{I}_k(t)\right]$$

*(Eq.18)*

Finally, we split the number of events into new recoveries and new fatalities as seen in equations 13-14.

#### 2.3. Environmental dependant reproduction number in livestock

<u>In this section we present how each environmental driver is accounted for in the model. Data sourcing and processing of environmental driver variables are available in section 4 of this document.</u>

##### Temperature-dependent reproduction number in livestock

Temperature is a known driver of viral transmission as it is a main factor driving *Hyalomma spp*. Adult *Hyalomma spp*. activity is known to occur above 12°C[1,2] and increases as temperature increases. Once temperature reaches above 30°C, ticks prefer to bury into soil[3]. The force of infection amongst livestock *λL(t)* (*Eq.8,9*) was therefore modelled as a linear function of temperature from 12 to 30$^{O}$ and linearly declining force of infection at temperatures above 30$^{O}$.

$$R_L(t) = \begin{cases} 0 & , if T < 12°C \\ A(T(t) - T_{min}) & , if 12°C \leq T \leq 30°C \\ A(30 - (T(t) - 30) - T_{min}) & , if T > 30°C \end{cases}$$

*(Eq.19)*

Where *A* is the temperature dependent transmission factor, and *T(t)* is soil temperature in °$C$ for Herat at time *t* of run-time (from ERA5 data).

##### Saturation-deficit-dependent reproduction number in livestock

Saturation deficit, a measure of the drying capacity of the air, is an implicit measure of air temperature. To keep tick activity within temperature-define range (12 -30 degrees C), we fit a polynomial function and predict the values and then find the corresponding saturation deficit value for the lower and max temperature values that define tick activity (Figure S3).

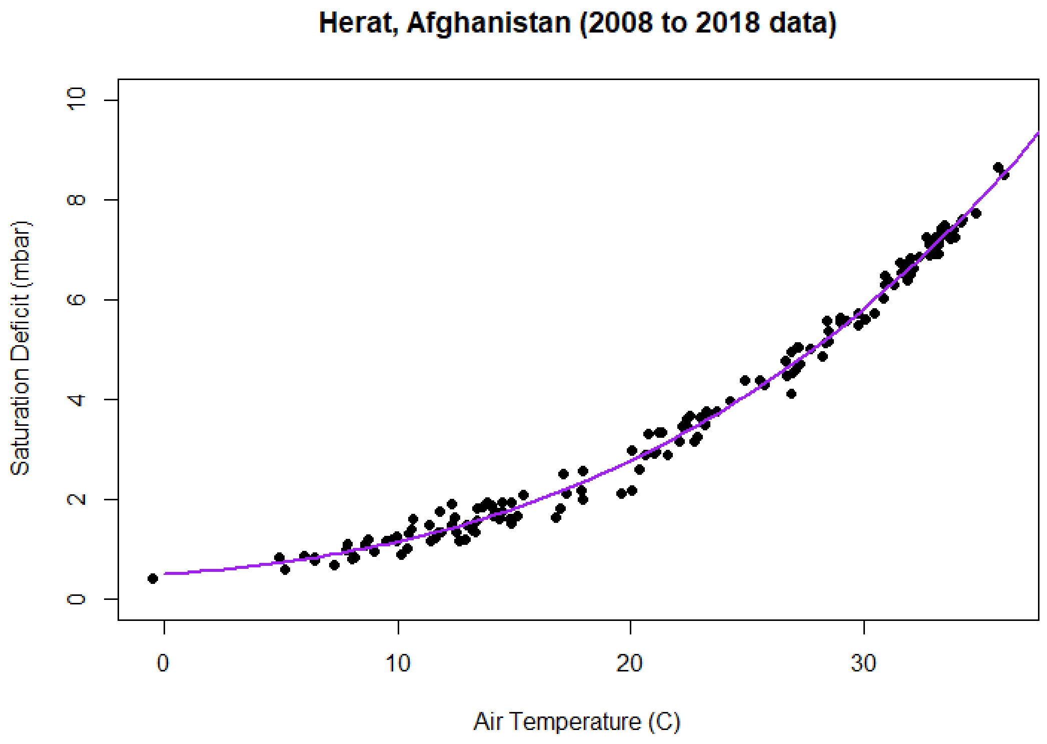

Figure S3. Polynomial model prediction model of Saturation deficit on Air temperature.

Therefore, we describe the saturation deficit dependent reproduction number as follows:

$$R_L(t) = \begin{cases} 0 & , if satdef < 6.415 mbar \\ A[min(satdef, 26.79mbar) - min\,(satdef)] & , if 26.79mbar \geq satdef \geq 6.415mbar \\ A[26.79mbar - (satdef - 26.79mbar) - min\,(satdef)] & , if satdef > 26.79mbar \end{cases}$$

*(Eq.20)*

Where *A* is here the saturation deficit dependent transmission factor, and *satdef(t)* is saturation deficit for Herat at time *t* of run-time (from ERA5 data).

#### Reproduction number in livestock for other environmental drivers

For the non-temperature drivers we define a force of infection as a linear function of the driver and the calibrated factor A. We don't impose limitations as caps or thresholds for these other drivers, namely, relative humidity and Normalized Difference Vegetation Index (NDVI). $R_L(t)$ is then defined as follows:

$$R_L(t) = A[driver(t) - min\,(driver)]$$

*(Eq.21)*

Where *driver* is to reflect the environmental factor used, either NDVI or relative humidity.

### 3. Livestock demographic model

The livestock model as explained above, is stratified into 5 age groups reflecting one year range for each group, with the last categories being those 4+ years. In order to obtain an equilibrium demographic model for livestock we simultaneously estimated the mortality rates for each age group. For this we constructed an age stratified births and deaths model, and optimised mortality rates for each age group to match a known livestock age distribution from a survey in Mauritania[4] using a simple optimisation algorithm in R (*bmle library*). In the absence of Afghani data on livestock age structure we choose data from Mauritania, a country with extensive livestock production for which this information has been published. We adapt the age distribution in livestock from Schulz et al [4], to reflect a mean life expectancy of 5 years. The total livestock population was obtained from FAO's 2003 survey[5]. Figure S4 shows the resulting age distribution.

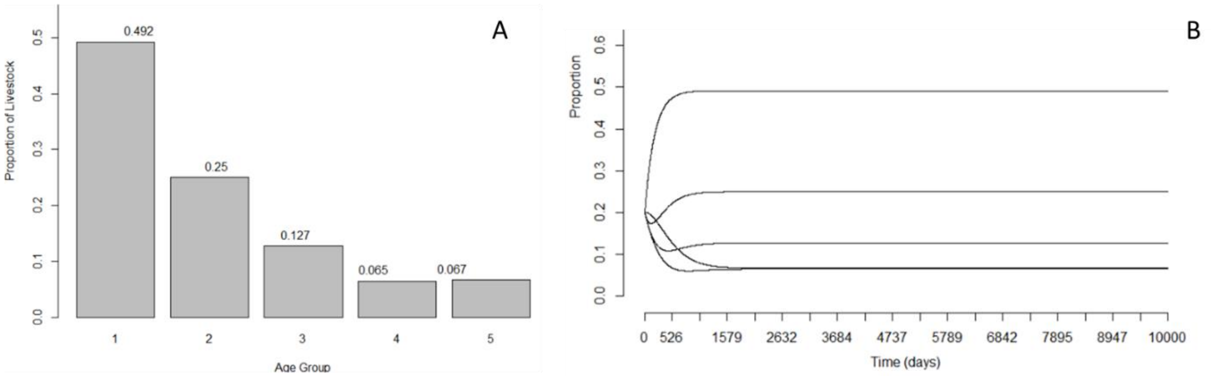

Figure S4. In panel A, a final age distribution in cattle into 5 age yearly age groups. In panel B, the time trend of age group proportions as they reach equilibrium.

### 4. Environmental drivers

In the absence of tick activity data we use (and test) environmental drivers as a surrogate measure of tick activity. It has been observed that *Hyalomma spp*. similarly to other tick species, are strongly driven in their reproduction cycles and also feeding activity by different climatic and land composition variables. Some of these variables are easily summarised by atmospheric indicators (soil temperature, relative humidity) and others require some calculation or a more complex approach (Saturation deficit , NDVI).

#### 4.1. Environmental data and indicator construction

Atmospheric data was retrieved for Herat, Afghanistan, ERA5 climate data reanalysis, and accessed through Copernicus project interface [6,7]. This dataset contains time series of multiple climatic variables around the planet.
We set up a polygon around Herat with Lat/Long coordinates (35°Lat / 62.1°Long - 34°Lat /62.5°Long).

For a description of drivers and its application in the model refer to table 2 in the main text.

<u>Soil temperature</u>

Retrieved for the selected area from April 2008 to Jan 2019. The time series of average monthly soil temperature was use directly as input into the model.

Relative Humidity

Relative humidity (RH) is not directly collected by the ERA5 project, but can be estimated using readily available variables like the 2m air temperature (airT2m) and the dewpoint temperature(dewpoint).

$$RH = \frac{actualvapourpressure}{saturatedvapourpressure}$$

(Eq.22)

$$actualvapourpressure = \frac{exp\left(17.625(dewpointT)\right)}{(243.04 + dewpointT)}$$

(Eq.23)

$$saturatedvapourpressure = \frac{exp\left(17.625(airT2m)\right)}{(243.04 + airT2m)}$$

(Eq.24)

Saturation deficit

Saturation deficit (SD) is not directly collected by the ERA5 project, but can be constructed from relative humidity (RH) and the ERA5 2m air temperature (airT2m)[8]

$$saturationdeficit = (1 - RH)(4.9463)exp\left(0.0621(airT2m)\right)$$

(Eq.25)

Normalized Difference Vegetation Index (NDVI)

NDVI is an index for the density of vegetation in a specific area. It is calculated by assessing the reflectance of vegetation as estimated with satellite images. NASA's Earth-Data project collates this information and estimates time series of NDVI, which we used and gathered from the MODIs subsets[9]. We use the same polygon of area around Herat described above. The time series data of monthly NDVI measures for the area is used directly into the model.

#### 5. Model calibration

We denote by $\theta$ the vector of input parameters, for all model inputs subject to uncertainty. For a given parameter set $\theta$, we followed the steps:

a. Run an instance of the Livestock model
b. Record Livestock output including a vector of the prevalence of infectious livestock at each time point.
c. Run an instance of the human spillover model with the relevant $\theta$ and the vector of prevalent I in livestock (to constrict force of infection, see Eq. 19-20)
d. Record human spillover model output
e. Calculate global posterior for the model and calibration targets described in table S1.

To compare these model projections with data *D*, we defined the posterior density $\pi(\theta)$ as:

$$\pi(\theta) \propto L(D \mid \theta).P(\theta)$$

*(Eq.26)*

Where $L$ is the likelihood of the data $D$ given models with parameters $\theta$ and $P$ is the joint prior distribution for $\theta$. For $P$, we took independent uniform distributions over the ranges shown in table 1 in the main text. The likelihood $L$ was constructed as follows. For count outputs (i.e., cases, fatalities) we estimate the *Poisson* likelihood, and the binomial likelihood for binary outcomes like prevalence. In particular, we determined the mean and variance of these distributions in order for the 2.5th, 50th and 97.5th percentiles to match respectively the lower, mid and upper ranges of estimates. For a given parameter set θ, we then constructed the overall likelihood π(θ) as a product of these distributions over all calibration targets listed in table S1. In practice we computed the logarithm of π(θ), thus taking the sum of the logarithms of each of the probability densities involved.

With $\pi(\theta)$ thus defined, we sampled the posterior density using a Markov Chain Monte Carlo approach. In brief, this approach implements a random walk through the space of parameter values $\theta$ to obtain an unbiased sample of the posterior density. We implemented the Metropolis Hastings algorithm with adaptive gaussian proposal existing in the package "fitR" for R [10]. For the set of parameter values thus obtained, we took every tenth element to reduce autocorrelation, thus yielding an 'ensemble' of parameters $\theta_1,\theta_2,\ldots$ ; This ensemble captures simultaneously the uncertainty in the parameter inputs, as well as in the calibration data. Then, to estimate uncertainty in a given simulated output ϕ (e.g. in the reduction of incidence with a given coverage of intervention), we simulated this output $\varphi_i$ for every $\theta_i$. We finally estimated uncertainty in $\varphi_i$ by determining its 2.5th, 50th and 97.5th percentiles. For graphic depiction of model fits to data see figure 2 in the main text 5. **Figure S5** shows the MCMC diagnostic outputs for each parameters, and **Figure S6** shows density plots for the calibrated parameters for the Saturation deficit model.

Convergence was assessed visually by inspecting the trace plots of the calibrated parameters and also through estimation of the Gelman –Rubin convergence diagnostic [11], computed as follows:

$$\ddot{R} = \frac{\ddot{v}}{w}, \qquad \textit{(Eq.27)}$$

Where $\ddot{v}$ is the posterior variance estimate of the combined chains and $w$ is the within-chain variance. If the chains have converged to the target posterior distribution, then $\ddot{R}$ (also known as the **scale reduction factor**) should be close to 1. As a rule of thumb, values below 1.1 are typically considered to indicate convergence. **Figure S7**, shoes the estimate of the Potential scale reduction factor (PSRF) for the six calibrated parameters.

| Calibration target | Description | Year(s) | Source |
|---|---|---|---|
| Livestock seroprevalence of CCHFV | Age stratified IgG seroprevalence from a serosurvey in Herat (n=132) | 2009 | Mustafa *et al*. 2011[12] |
| Human seroprevalence of CCHFV | IgG seroprevalence I humans by occupation in Herat (n=330) | 2009 | Mustafa *et al*. 2011[12] |
| Monthly Human CCHFV cases reported | Reported human cases in Herat. Cases in 2018 at national level and assumed that ~62% are from Herat according to Niazi et al.[13] | 2008, 2017, 2018 | Mofleh et al[14]<br>Niazi et al[13]<br>Sahak et al[15] |
| Yearly Human CCHFV cases reported | Yearly aggregated cases reported nationally. Assumed that ~62% are from Herat according to Niazi et al.[13] | 2009, 2010, 2010, 2011, 2012, 2013, 2014, 2015, 2016 | Niazi et al[13]<br>Sahak et al[15] |
| Yearly Human CCHFV fatalities reported | Yearly aggregated deaths reported nationally. Assumed that ~62% are from Herat according to Niazi et al.[13] | 2009, 2010, 2010, 2011, 2012, 2013, 2014, 2015, 2016 | Niazi et al[13]<br>Sahak et al[15] |

**Table S1: Calibration target datasets for CCHFV in Herat, Afghanistan**

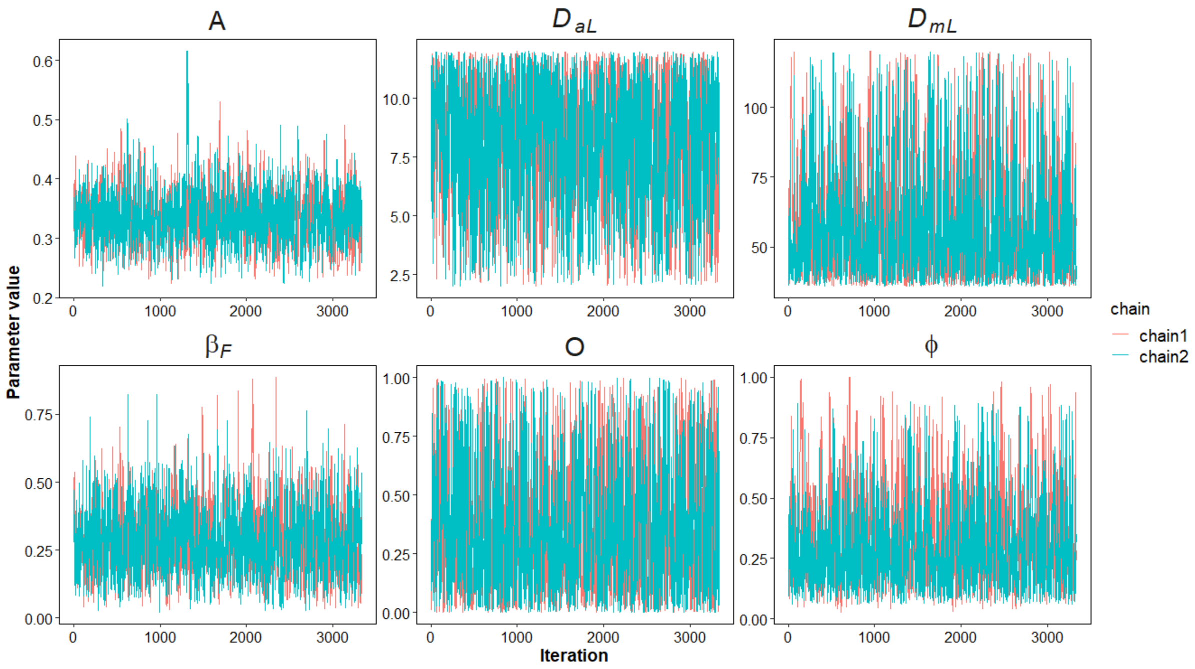

**Figure S5: MCMC Trace plots** for the 6 calibrated parameters. Plots show two 100K chains (after burn and thinning). These results reflect calibration for the final selected model (saturation deficit). Trace plots show a good mixing of the chains which suggests convergence to a stationary distribution.

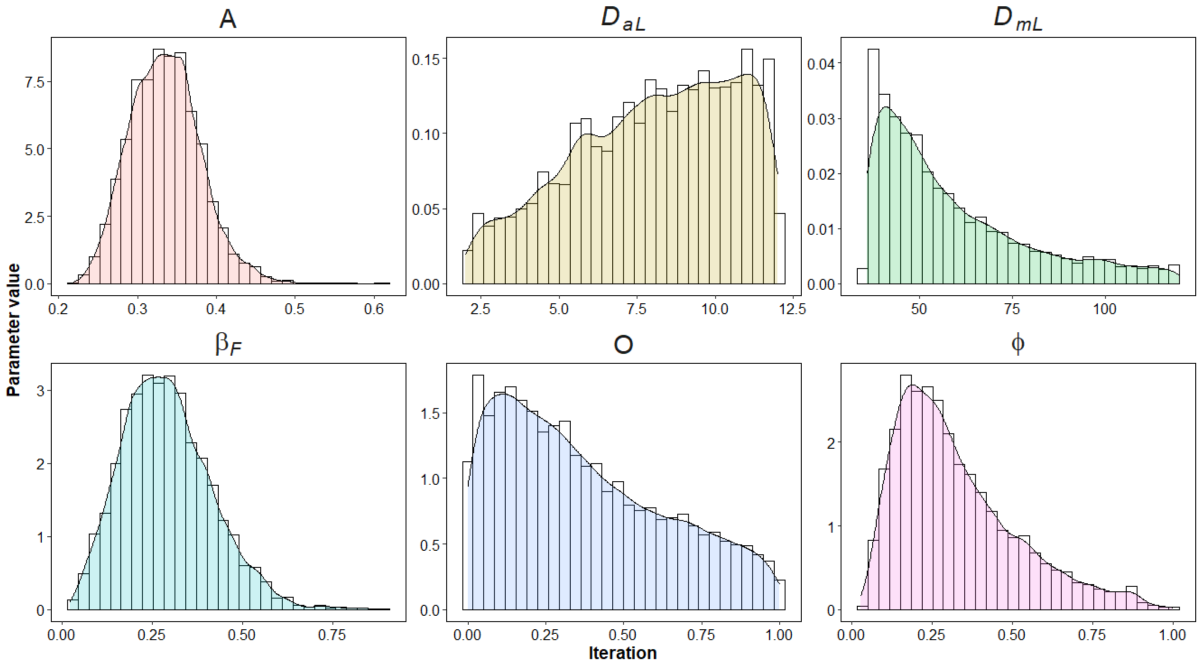

**Figure S6. Density plots.** Histograms and density plots sampled from the posterior distribution obtained through MCMC. Each parameter reflects the combined, burned, thinned and resampled posterior for the calibrated parameters. These are results for the final selected model, i.e., with saturation deficit as the environmental driver.

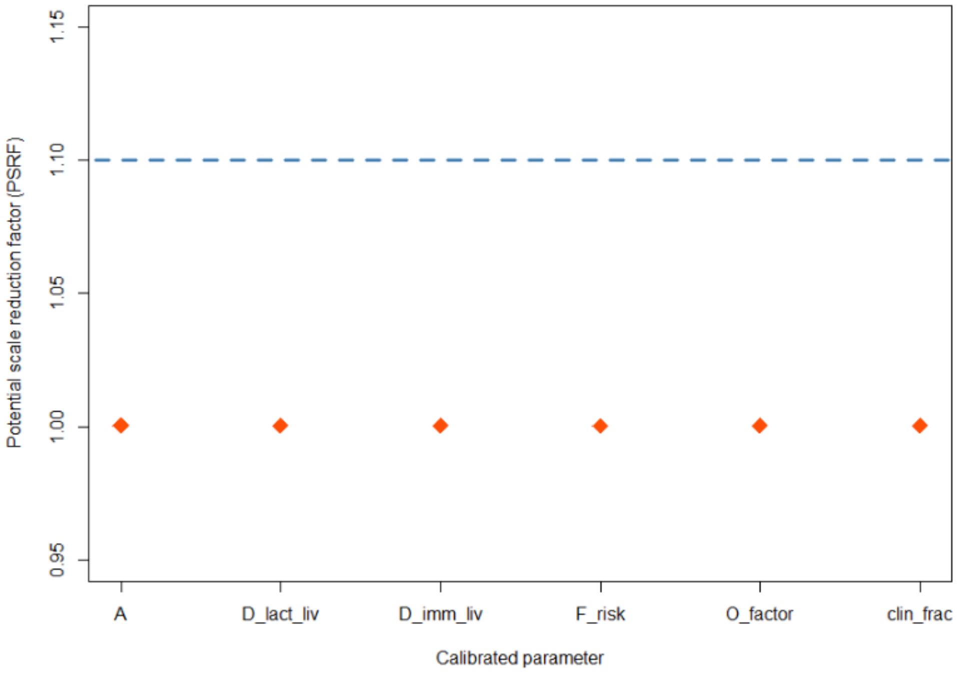

**Figure S7: Gelman-Rubin diagnostic** for the six calibrated parameters. Orange dots show the point estimate of the Potential Scale Reduction Factor for each calibrated parameter, and the dashed blue line shows the rule-of-thumb convergence threshold. This suggests good convergence for all parameters. These are results for the final selected model, i.e., “saturation deficit driver” model. Variations in specific PSRF are not evident here given the scale of the plot.

### 6. Further modelling analysis

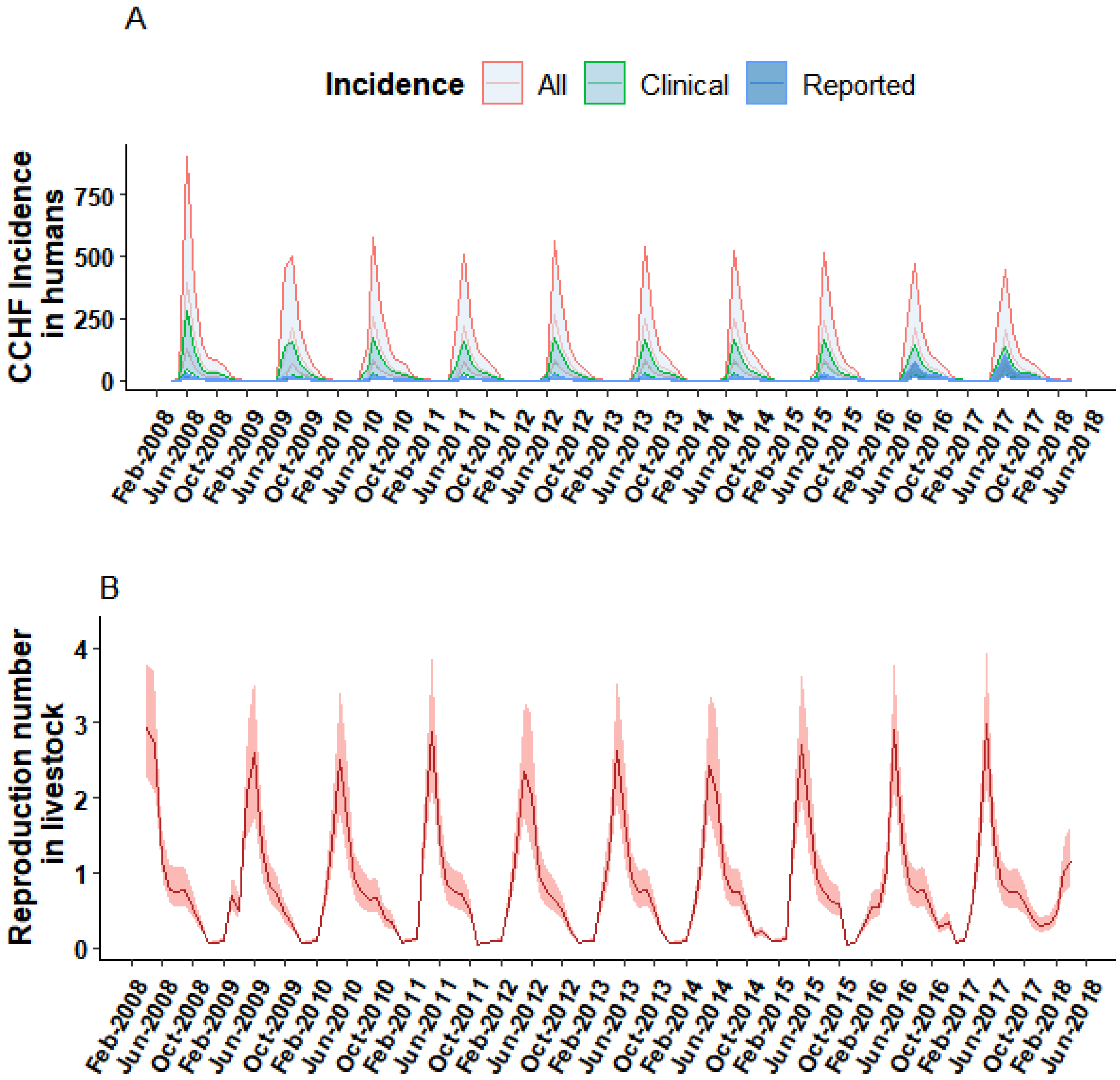

**Figure S8: Transmission dynamics of CCHF in Herat, Afghanistan 2008-2018**
In panel A, simulated trajectories of monthly CCHF incidence in a spectrum from reported clinical cases (blue shade), to all clinical cases (green) and all cases (red) including asymptomatic/subclinical cases. The shaded area shows the 95% CrI. In Panel B, the simulated effective reproduction number for CCHFV in livestock. These are results for the final selected model, i.e., “saturation deficit driver” model.
